# Combined High-Resolution MRSI and [18F]-FACBC PET to Improve the Presurgical Diagnostic Accuracy in Gliomas

**DOI:** 10.1101/2025.10.01.25336990

**Authors:** Erin Beate Bjørkeli, Anna Maria Karlberg, Benedikte Emilie Vindstad, Lars Kjeldsberg Pedersen, Ole Skeidsvoll Solheim, Jonn Terje Geitung, Morteza Esmaeili, Live Eikenes

**Affiliations:** Department of Diagnostic Imaging, Akershus University Hospital, Lørenskog, Norway; Institute of Clinical Medicine, University of Oslo, Oslo, Norway; Department of Circulation and Medical Imaging, Norwegian University of Science and Technology (NTNU), Trondheim, Norway; Department of Radiology and Nuclear Medicine, St. Olavs Hospital, Trondheim University Hospital, Trondheim, Norway; Department of Neurosurgery, Ophthalmology and Otorhinolaryngology, University Hospital of North Norway, Tromsø, Norway; Department of Neurosurgery, St. Olavs Hospital, Trondheim University Hospital, Trondheim, Norway; Department of Neuroscience, Norwegian University of Science and Technology (NTNU), Trondheim, Norway; Department of Electrical Engineering and Computer Science, University of Stavanger, Stavanger, Norway

**Keywords:** Deep learning, Glioblastoma, neuro-oncology, Fluciclovine PET Molecular imaging, Multimodal imaging, Metabolite MRI, Metabolite maps

## Abstract

Medical imaging is crucial for glioma management. Combined with MRI, amino acid PET may improve glioma diagnosis, biopsy targeting, and tumor delineation compared to structural MRI alone. Magnetic resonance spectroscopic imaging (MRSI) complements both structural MRI and PET by detecting metabolites such as N-acetylaspartate (NAA), creatine (Cr), and choline (Cho), which are markers for brain health and tumor malignancy, but is challenged by low spatial resolution. This study evaluates whether high-resolution MRSI enhanced by deep learning can improve diagnostic accuracy and serve as a complement or alternative to PET for glioma classification. Ten glioma patients (CNS WHO grades 2–4, ages 24–80) were included. Presurgical [18F]-FACBC PET/MRI, including proton 2D MRSI, was acquired for all patients. Thirty image-guided biopsies were sampled during surgery from the patients and classified as glioma tissue or non-tumor tissue, and according to IDH1 status. For each biopsy location, tumor-to-background ratio (TBR) and standardized uptake value (SUV) from PET, and tCho/NAA and tCho/tCr ratios from MRSI were calculated. ROC analysis was used to assess the accuracy of [18F]-FACBC PET and high-resolution MRSI, and the combinations of these in classifying glioma vs. non-tumor tissue and IDH1 status. The tCho/NAA ratio from the deep learning-based model demonstrated excellent diagnostic accuracy in classifying glioma vs. non-tumor tissue (AUC = 0.87, 95% CI: 0.66– 1.0), outperforming SUV (AUC = 0.71, 95% CI: 0.49–0.90), TBR (AUC = 0.68, 95% CI: 0.48–0.86), and tCho/tCr (AUC = 0.81, 95% CI: 0.54–1.00). Combining TBR with tCho/NAA and/or tCho/tCr improved tissue classification compared to either modality alone, where TBR + tCho/NAA + tCr/NAA showed the best results (AUC = 0.91, 95% CI: 0.71–1.0). MRSI was a poor predictor for IDH1-status (tCho/NAA: AUC = 0.67, 95% CI: 0.44–0.88 and tCho/tCr: AUC = 0.38, 95% CI: 0.17–0.60), while PET was an excellent predictor (SUV: AUC = 0.83, 95% CI: 0.66–0.85 and TBR: AUC = 0.82, 95% CI: 0.65–0.94) and the combination of SUV and tCho/tCr was an outstanding predictor (AUC = 0.96, 95% CI: 0.88–1.0). Incorporating high-resolution MRSI in combination with [18F]-FACBC PET improved the diagnostic accuracy in differentiating glioma tissue from non-tumor tissue.

**Significance Statement:** Our study highlights the importance of combining imaging methods for brain tumor characterization. MRI remains central in brain imaging but is limited, making PET a valuable molecular complement. MRSI provides insight into neurometabolic alterations associated with tumor growth, yet its clinical utility has been limited by low spatial resolution. By applying deep learning, we enhanced the resolution of MRSI and compared its performance with PET. Our findings demonstrate that High-resolution MRSI adds diagnostic value and, with PET, may enhance glioma classification and inform future clinical decision-making.

---

**A**dult diffuse gliomas are classified according to the 2021 World Health Organization (WHO) classification of tumors of the central nervous system (CNS) into glioblastoma *IDH* -wild-type (grade 4), astrocytoma *IDH* -mutant (grades 2–4), and oligodendroglioma *IDH* -mutant with 1p/19q co-deletion (grades 2–3) (1). Gliomas are highly heterogeneous, with regions displaying different properties and degrees of malignancy. Presurgical imaging is important for tumor delineation, predicting diagnoses, and surgical planning (2, 3).

Structural magnetic resonance imaging (MRI) is the imaging method of choice for brain tumor examinations due to its high-resolution soft tissue contrast, but may have limitations when it comes to identifying the most malignant regions. Metabolite imaging is a group of diagnostic techniques that provide non-invasive visualization of metabolic activity in living tissues and can be visualized by either positron emission tomography (PET) or magnetic resonance spectroscopy imaging (MRSI). Alterations in metabolic activity can be indicative of underlying pathologies and may exhibit both the presence and aggressiveness of tumors (4). Metabolic imaging can therefore aid in identifying the most aggressive regions of the tumor. MRSI and PET are often used in combination with MRI techniques to complement anatomical imaging.

Glioma cells show increased transmembrane amino acid transport to facilitate enhanced protein synthesis. The amino acid PET tracer 18F-fluorocyclobutane-1-carboxylic acid ([18F]-FACBC) takes advantage of this mechanism, utilizing amino acid transporters for cellular uptake. Previous studies using [18F]-FACBC PET in gliomas demonstrated its utility in distinguishing tumors from normal brain tissue and correlating uptake patterns with histopathological findings. In comparison with MRI performed according to recognized standards for examining gliomas, [18F]-FACBC PET improved diagnostic accuracy in identifying tumor infiltration beyond visible abnormalities on structural MRI (5–8). This highlights its potential role in guiding neurosurgery and therapeutic planning by delineating metabolically active tumor regions more effectively than structural MRI.

Complementary to this, MRSI provides information on the tissue content of specific metabolites, such as N-acetyl aspartate (NAA), creatine (Cr), and choline-containing metabolites (tCho) (9, 10). These metabolites are present in lower concentrations compared to the dominant water and lipid signals, typically in the range of a few millimoles per liter (mM) within brain tissues (11). Concentrations may vary in different regions of the brain and can also be influenced by factors such as age, health status, and metabolic activity (12–14). In gliomas, decreased NAA indicates neuronal loss, whereas elevated tCho reflects increased membrane turnover; thus, elevated tCho/tCr and tCho/NAA ratios serve as key biomarkers for tumor detection (15) and treatment evaluations (16). These variations make metabolites and their ratios valuable markers for brain examinations. However, MRSI is limited by its lower spatial resolution compared to structural MRI (normally 1 mm isotropic resolution). Signal averaging or the use of larger voxel volumes is essential to enhance the signal-to-noise ratio (SNR) in MRSI (17). However, increasing voxel size reduces spatial resolution in MRSI sequences, and typically the spatial resolution is limited from 16 × 16 to 64 × 64 voxels. For instance, a standard 32 × 32 matrix covering the adult brain with a field-of-view of 220 mm^2^ yields a nominal voxel size of approximately 6.875 × 6.875 mm^2^. Even with advanced research protocols, spatial resolution of MRSI remains coarser than other MR modalities. Post-processing techniques have been developed for high-resolution MRSI, but many of these methods encounter challenges such as high complexity and suboptimal results. Deep learning models have, in recent years, been tested as alternative upscaling methods, with promising results (18–20).

This study aimed to investigate the diagnostic accuracy of high-resolution MRSI, both compared to and combined with [18F]-FACBC PET, for classifying glioma versus non-tumor tissue and to determine IDH1 status in patients with gliomas, using image-guided biopsies as reference. The image quality of the low-spatial resolution MRSI data was increased using a deep learning-based model trained to upscale lower-resolution metabolite maps.

## Results

### Biopsy Coverage

Although DL-based maps still covered less part of the tumor and therefore fewer biopsies than PET (13.5% of total biopsies are not covered by DL-based maps, *p* =0.06), they significantly improved coverage compared to the nearest-neighbor method (23.5%, *p* =0.005) (**Fig. 1**).

**Fig. 1.**
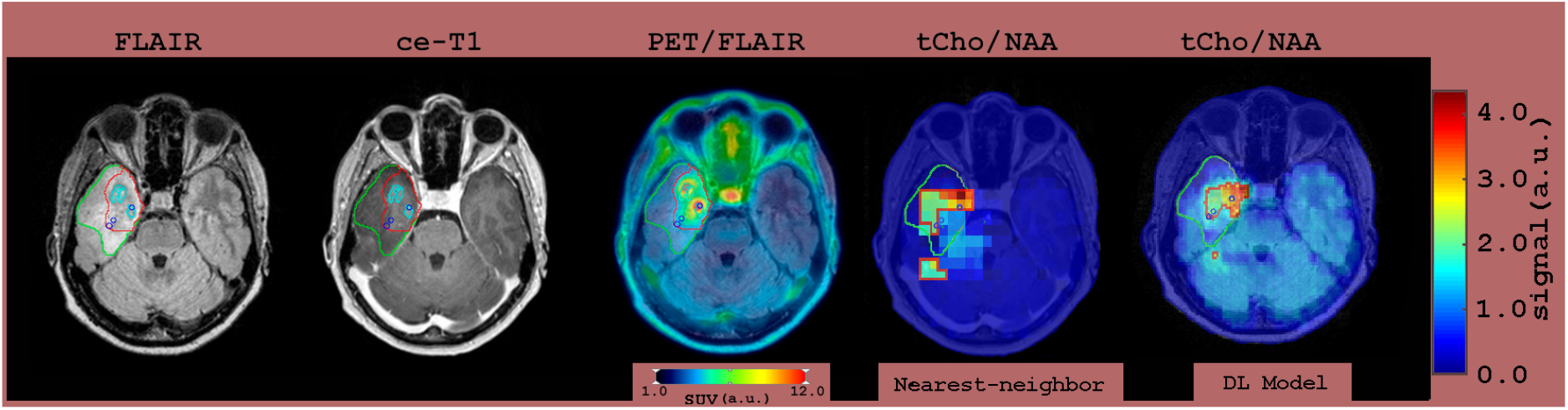
An example axial view from (left to right): fluid-attenuated inversion recovery (FLAIR), contrast-enhanced T1-weighted (ce-T1), combined FLAIR/PET, and upscaled total-Choline/N-acetyl aspartate (tCho/NAA) maps using nearest-neighbor interpolation and the DL-model for a patient with glioma. Colored contours indicate tumor area/volume determined by FLAIR (green), ce-T1 (cyan), TBR *≥* 2.0 (red, PET image), and tCho/NAA *≥* 2.37 (red). Blue circles indicate biopsy sites classified as high-grade glioma. Notably, two biopsies lacked contrast enhancement but exhibited [18F]-FACBC uptake *≥* 2.0. One biopsy is outside the margin determined by the tCho/NAA map upscaled using the nearest-neighbor method, while all biopsies (blue circles, rightmost panel) are included within the tumor area/volume determined using DL model upscaling.

### Classification of Glioma vs. Non-tumor Tissue

In the ROC analysis, the DL-based MRSI provided significantly higher diagnostic accuracy with an AUC of 0.87 (95% CI: 0.66-1.00) and 0.81 (95% CI: 0.54-1.00), compared to the nearest-neighbor interpolation demonstrating poor diagnostic accuracy with AUC of 0.55 (95% CI: 0.26-0.82) and 0.62 (95% CI: 0.36-0.87) for tCho/NAA and tCho/tCr, respectively (*p <*0.02). Further results are therefore presented only for the DL-based MRSI maps. The results from the ROC analysis of glioma/non-tumor classification of the biopsies are presented in **Figure 2**. The performance of the tCho/NAA and tCho/tCr ratios is shown in **Figure 2**, demonstrating their distinct diagnostic accuracy. However, tCho/tCr exhibited the highest sensitivity (0.92). SUV had acceptable performance (AUC =0.71, 95% CI: 0.49-0.90) and TBR had poor performance (AUC =0.68, 95% CI: 0.46-0.86). All values are listed in the **Table S1**.

**Fig. 2.**
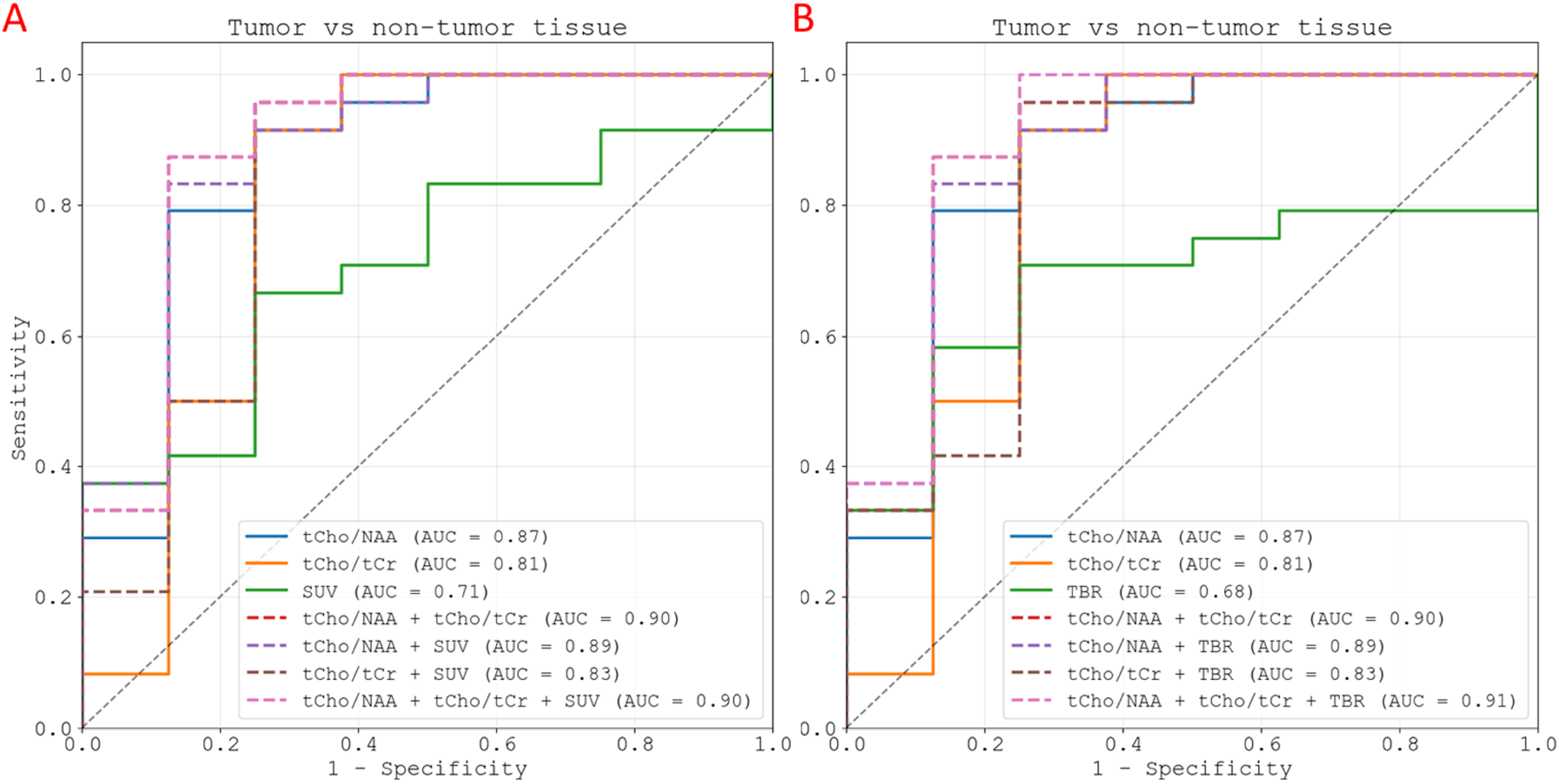
Receiver operating characteristic (ROC) curve analysis was performed as a measure for the diagnostic accuracy of glioma vs. non-tumor tissue classification for tCho/NAA, tCho/tCr, SUV (**A**), TBR (**B**), and a combination of all covariates in the discrimination of glioma from non-tumor tissues. The MRSI-derived metabolite ratios are from the deep-learning upscaled MRSI maps.

The combination of TBR or SUV with tCho/NAA or tCho/tCr improved the glioma/non-tumor classification compared to either modality alone, as demonstrated with the ROC-curves in **Figure 2** (**Table S1**) and the confusion matrix in **Figure 3**. The optimal combination, tCho/NAA + tCho/tCr + TBR, achieved an AUC of 0.91 (95% CI: 0.71-1.00). However, the heatmap of the DeLong analysis in **Figure 4** shows that only the combinations including TBR, tCho/NAA and/or tCho/Cr provide significantly higher diagnostic accuracy than the individual modalities (*p <*0.05, log10(*p*) *>*1.3).

**Fig. 3.**
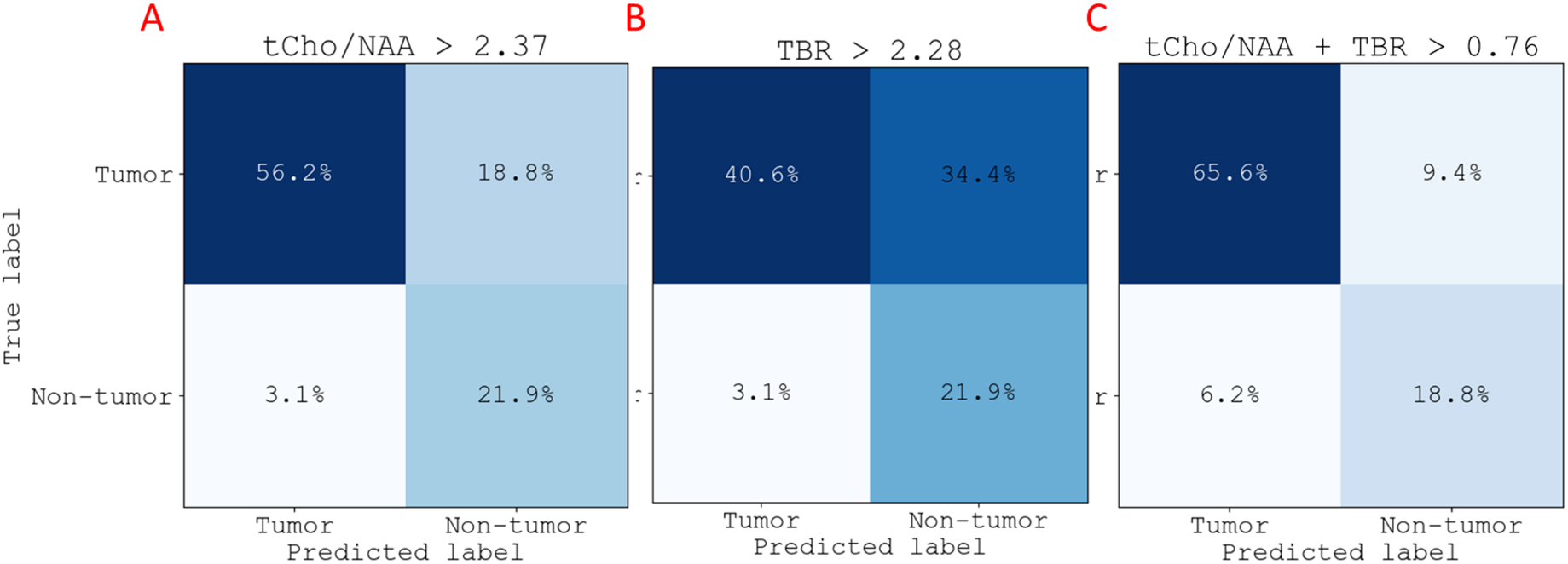
Confusion matrix using the optimal threshold from ROC-analysis for (**A**) tCho/NAA, (**B**) TBR, and (**C**) the combined modality of TBR and tCho/NAA for discrimination of glioma vs. non-tumor tissue classification, than either modality alone. MRSI-derived metabolic ratios are calculated using the deep-learning upscaled MRSI maps.

**Fig. 4.**
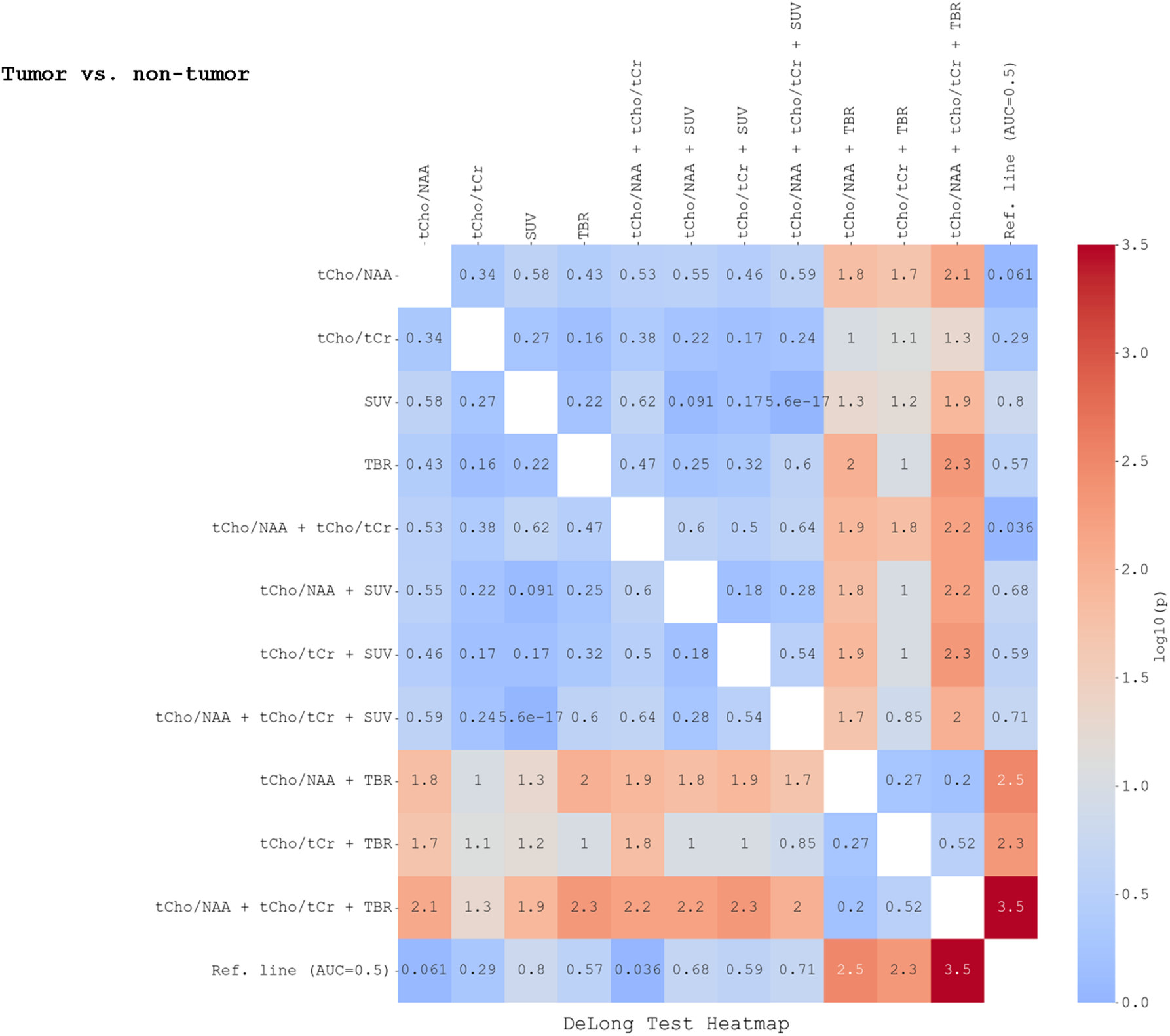
Heatmap showing the result of the DeLong test to evaluate statistical significance among ROC curves assessing the diagnostic accuracy of glioma vs. non-tumor tissue classification (curves for tCho/NAA, tCho/tCr, SUV, TBR, and the combination of them). Results are listed as log10 (*p*-values), with values *>*1.3 (light orange to dark red) indicating statistical significance. MRSI-derived metabolic ratios are calculated using the deep learning-upscaled MRSI maps.

### Classification of IDH1 Mutation Status

The results from the ROC analysis of IDH1 mutation status of the biopsies are presented in **Figure 5**. The metabolite ratios tCho/NAA and tCho/tCr show poor performance for classifying IDH1-mutation status (AUC=0.67, 95% CI: 0.44-0.88 and AUC =0.38, 95% CI: 0.17-0.60). TBR and SUV both demonstrated stronger performance, with SUV being slightly better with an AUC of 0.83 (95% CI: 0.66-0.95) for IDH1 status. The combination of SUV, tCho/NAA, and tCho/tCr improved accuracy for classification of IDH1 mutation status (AUC =0.96, 95% CI: 0.88-1.00, *p <*0.05) compared to standalone metabolite ratios (**Figs. 5-6, Table S2**).

**Fig. 5.**
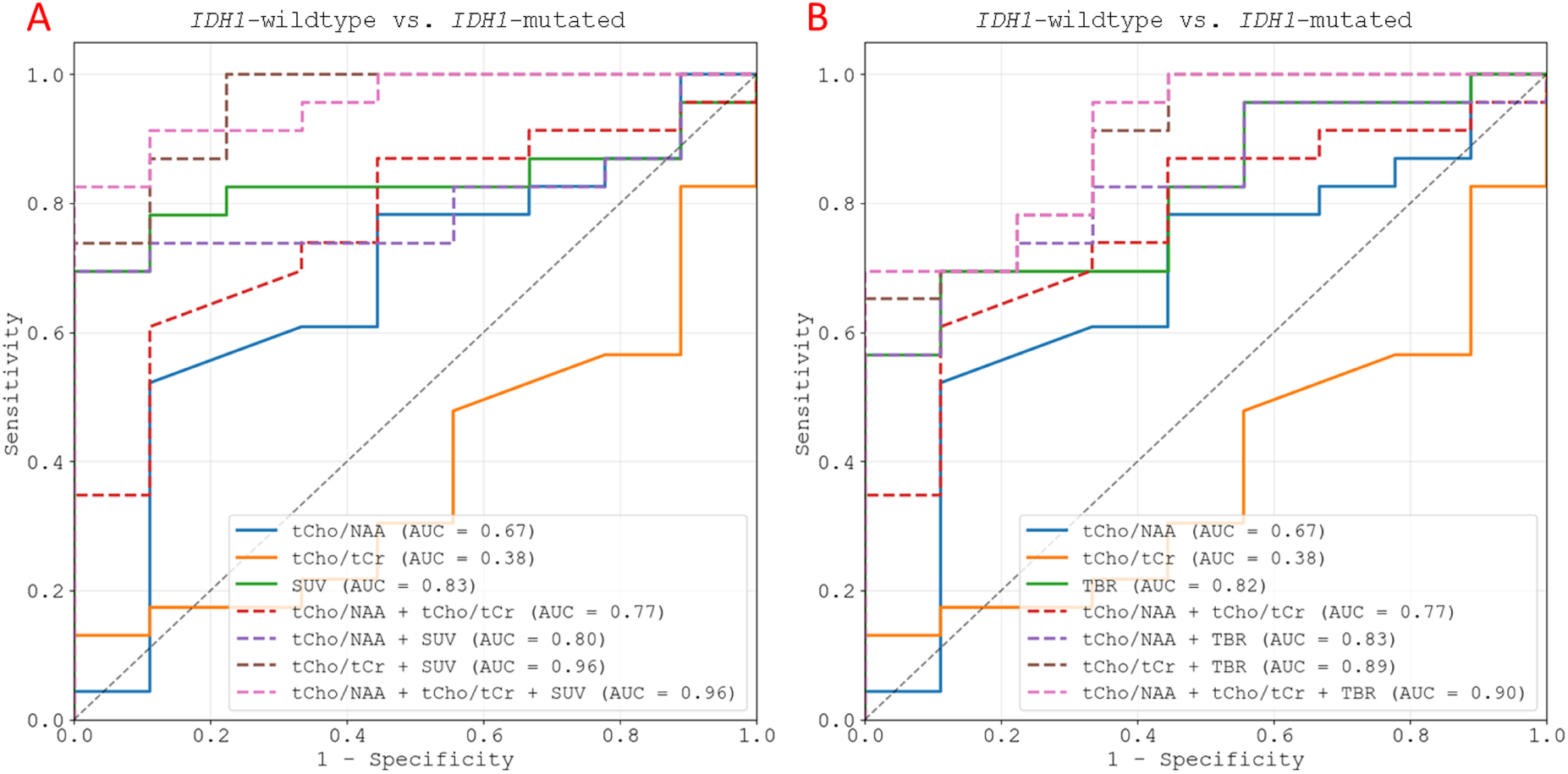
Receiver operating characteristic (ROC) curves for MRSI-derived metabolite ratios (tCho/NAA, tCho/tCr) and SUV (**A**) or TBR (**B**), and a selective combination of covariates when applied to discrimination of *IDH1*-wildtype from *IDH1*-mutated tissue. MRSI-derived metabolic ratios are calculated using the deep learning-upscaled MRSI maps.

**Fig. 6.**
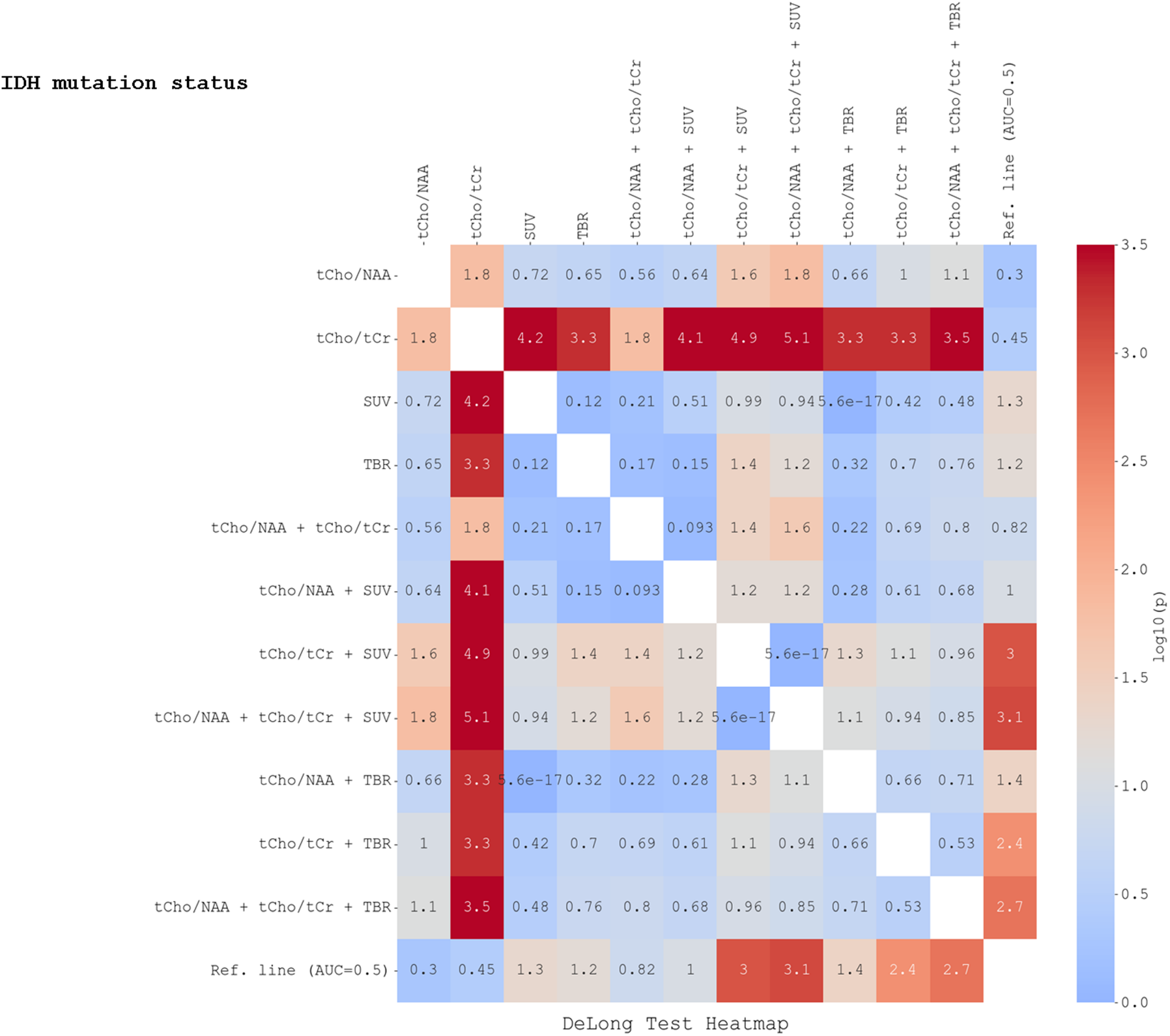
Heatmap showing the result of the DeLong test to evaluate statistical significance between the ROC curves for IDH1-mutation status. Results are listed as log10 (*p*-values), with values *>*1.3 (light orange to dark red) indicating statistical significance. MRSI-derived metabolic ratios are calculated using the deep learning-upscaled MRSI maps.

## Discussion

The results presented in this study highlight the diagnostic potential of metabolic ratios derived from MRSI and suggest that tCho/NAA and tCho/tCr can improve diagnostic accuracy beyond [18F]-FACBC PET alone. A combination of TBR, tCho/NAA, and tCho/tCr yielded the highest AUC (AUC =0.91, *p <*0.05) for classification of glioma tissue vesus non-tumor tissue, while combining tCho/NAA, tCho/tCr, and SUV improved the classification of *IDH1* -wildtype and *IDH1* -mutated tissues (AUC =0.96, *p <*0.05) compared to the standalone metabolite ratios.

MRSI did not cover the entire tumor and was not applicable for calculation in all biopsies. In this study, two-dimensional MRSI slices were selectively placed in a region covering a larger area of the lesion, successfully identifying parts of the tumor in most subjects. The time window for MRSI examination is extremely limited when acquired after the other MR sequences, such as fluid-attenuated inversion recovery (FLAIR), T1W (with/without contrast), T2W, DWI, perfusion, and potentially functional MRI. Thus, the choice of slab MRSI acquisition over whole-brain MRSI was due to the longer scan time of the vendor-provided 3D PRESS sequence, which is approximately 20 minutes (20). The use of recently developed faster 3D MRSI techniques, with scan times under seven minutes, enables whole-brain metabolite imaging, allowing comprehensive lesion coverage (16, 21, 22).

An optimal outcome of the upscaling method would be an improved effective spatial resolution, enabling more detailed visualization of structures, particularly in areas that are typically difficult to distinguish, such as edges (23–25). The DL method improves the effective spatial resolution markedly by learning to fill in more accurate and detailed information, especially around edges and boundaries (23), which would be missed by traditional methods such as nearest-neighbor and spline interpolation. The improved performance of the DL method may be attributed to the incorporation of prior anatomical knowledge, allowing for more precise metabolic mapping and increased alignment with tumor regions, especially in complex, heterogeneous gliomas, as demonstrated in previous studies (19, 20). Given the limited number of patients in this study, we cannot yet conclude the utility of the DL method in the precise diagnosis of gliomas, but our results suggest its potential.

Compared to normal healthy brain tissue, increased levels of tCho/NAA and tCho/tCr have been previously reported in malignant gliomas (15, 16, 26, 27). In this study, tCho/NAA was the most accurate modality, with an AUC of 0.87 in differentiating glioma from non-tumor tissue. Despite this, tCho/tCr had the highest sensitivity (0.92), suggesting that this parameter might be a valuable diagnostic tool for detecting subtle metabolic changes, though with a lower specificity than tCho/NAA. TBR showed poor and SUV acceptable performance according to AUC measures, which may indicate limited diagnostic utility as standalone measures, at least in our cohort. However, it is important to acknowledge that comparisons between PET and MRSI are influenced by the imaging method used for biopsy selection, since PET and MRSI show different metabolic changes. Compared to our results, a previous study reported an opposite trend, where amino acid PET using [18F]-FET yielded higher AUC values than MRSI (28). In their study, due to the lack of MRSI data at the time of biopsy, trajectories were planned based on PET, targeting the most metabolically active tumor regions. This study design, therefore, inherently favored [18F]-FET PET over MRSI. However, previous studies (5, 26) have demonstrated the potential utility of MRSI in biopsy planning, suggesting that a prospective study incorporating both modalities could provide a more balanced and comprehensive comparison. Another major contributing factor for discrepancies in reported diagnostic performance is threshold determination for SUV, which varies across studies and is influenced by factors such as the amino PET-tracer used, PET scanner differences, reconstruction variations, and patient selection criteria. As previous research has shown, these parameters can cause slight shifts in threshold values, highlighting the need for larger, biopsy-controlled studies to establish and validate optimal cutoff values.

Gliomas are heterogeneous, with regions of varying activity and molecular properties, complicating diagnosis and the identification of aggressive tumor areas (3). Compared to PET/CT, MRSI combined with amino acid PET has been shown to enhance diagnostic accuracy for patients with gliomas by improving tumor detection, characterizing heterogeneity, and assessing treatment responses more effectively (29–33). Furthermore, a prospective study of 50 glioma patients by Floeth et al. (34) demonstrated that integrating [18F]-FET-PET and MRS significantly improved diagnostic accuracy. When both modalities were positive, histological confirmation of tumors reached 97%, whereas negative findings consistently indicated nonneoplastic disease. These results underscore the added value of PET and MRSI in cases where MRI alone is inconclusive (34). The increased diagnostic accuracy could be caused by the fact that the two methods can provide complementary information, as PET primarily reflects specific metabolic pathways, while MRSI provides biochemical concentrations within the tumor (35–37).

As amino acid PET imaging becomes more widely available, it is increasingly recognized as a reliable marker of glioma infiltration and IDH status. [18F]-FET PET has shown promise in distinguishing *IDH1* -wildtype from *IDH1* -mutated gliomas by leveraging metabolic differences, aiding diagnosis and treatment planning (38–42). Studies using TBR demonstrated their effectiveness in differentiating molecular subtypes, aligning with our findings and highlighting the value of amino acid PET-derived parameters like SUVs and TBRs for noninvasive IDH status characterization. However, none of these studies incorporated MRSI data, limiting direct comparison with our results on combined PET/MRSI modalities for identifying *IDH1* -mutated gliomas. Recent studies have demonstrated the utility of [18F]-FET PET (43) and emerging PET tracers (44, 45) targeting *IDH* mutations, suggesting a potential paradigm shift in the diagnosis of gliomas and treatment planning.

One limitation of the study is the relatively small sample size. Most of the tumors are of a higher grade, and the results of this study might not be transferable to low-grade samples and less common glioma types. There is also a lack of biopsies from normal tissue to be used as “control”, as biopsies were only targeting the tumors according to increased FLAIR signal. This might affect the calculation of specificity, due to an imbalance in the dataset. Furthermore, our deep learning model, which was used to upscale the MRSI data, was trained on synthetic datasets. This could limit its generalizability, as it does not account for several challenging factors, such as eddy current artifacts, field inhomogeneity, variable acquisitions, and MR system setups. Additionally, the inherent black-box nature of deep learning models raises concerns regarding ethical issues and risk management, due to the lack of interpretability, and by extension, the trustworthiness of their outputs (46, 47).

Future works may focus on addressing the study’s limitations, such as the small sample size, by expanding the cohort to include low-grade gliomas. Additionally, training deep learning models on in vivo datasets with varied artifacts and MR setups could enhance model generalizability. Exploring multi-modal imaging and improving the interpretability of AI models will also be important for addressing ethical concerns and improving clinical trust. These efforts could advance AI-powered glioma precision medicine.

## Conclusion

The deep learning-based MRSI model using the tCho/NAA ratio outperformed SUV, TBR, and tCho/tCr in diagnosing glioma versus non-tumor regions, while combining TBR with tCho/NAA and/or tCho/tCr improved tissue classification compared to either modality alone. For distinguishing IDH1-mutation status, SUV and TBR outperformed the MRSI-derived parameters, while a combination of tCho/tCr with TBR and SUV slightly improved the classification. Our results reinforce the notion that PET and MRSI provide complementary insights that can enhance diagnostic. Future studies should aim to refine the integrated PET/MRSI approach to optimize study design and assess their combined clinical impact in larger, biopsy-controlled cohorts. Standardizing acquisition protocols and threshold values across different PET tracers and scanners will also be crucial in improving reproducibility and reliability in glioma diagnostics.

## Materials and Methods

### Study Participants

Twenty-three adult patients undergoing primary resection for diffuse glioma underwent [18F]-FACBC PET/MRI, including MRSI. Among these, 13 underwent image-guided biopsies. The biopsies were taken from both PET-hotspot and PET-negative areas, when possible. All biopsies were sampled from areas with increased FLAIR signal (6). Three patients were excluded due to loss of image-localization data (n =1) or uncertain diagnosis based on biopsy (n =2), leaving 10 patients with a total of 30 image-guided biopsies for inclusion in the current study (**Table 1**). All participants gave written informed consent. This study was performed in accordance with the Declaration of Helsinki and approved by the Regional Ethics Committee (REK South East Norway, reference number: 2018/2243).

**Table 1.**
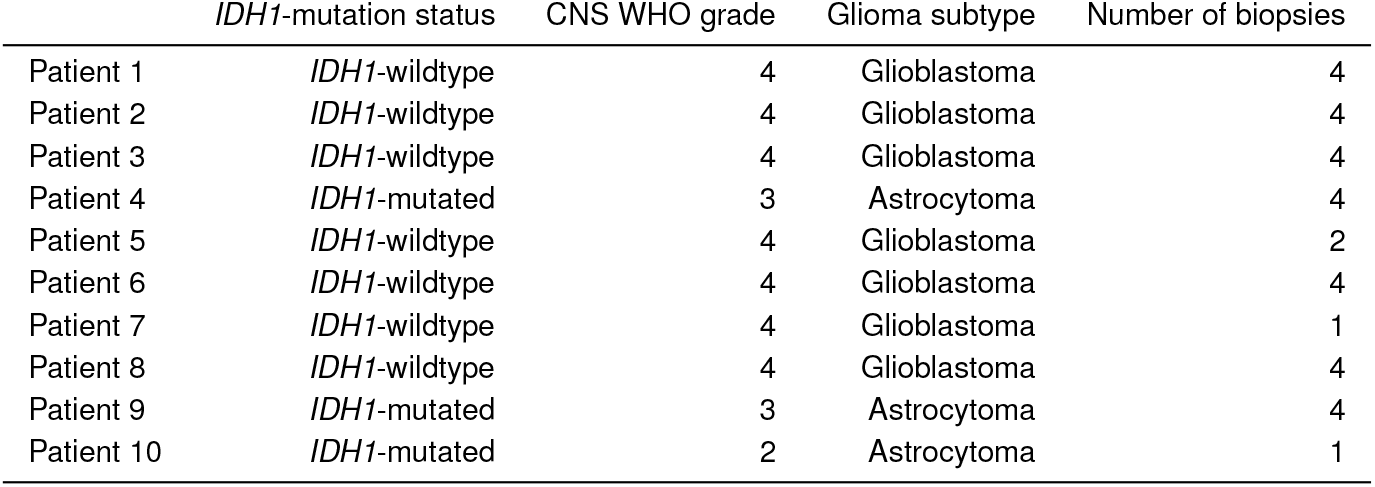
Summary of patient information.

### Biopsy Analysis

Twenty-four of the 30 biopsies were classified as glioma tissue, while six were classified as non-tumor tissue. Non-tumor tissue was defined as normal tissue and/or mild inflammatory changes, while necrotic tissue was classified as glioma tissue. The classification was performed by a neuropathologist and based on immunohistochemical analysis (n =11), DNA methylation analysis (n =10), or both (n =9). The biopsies were also classified as *IDH1* -wildtype (n =8) or *IDH1* -mutated (n =22). A more detailed description is provided by Vindstad et. al (6).

### PET/MRI Acquisition and Reconstruction

Anatomical MRI, proton 2-dimensional MRSI, and [18F]-FACBC PET were acquired using a hybrid PET/MRI system (Siemens Biograph mMR, software version Syngo MR, Erlangen, Germany). The patients received an injection of [18F]-FACBC (3 ± 0.2 MBq/kg) on the scanner examination table at the onset of the PET/MRI acquisition, and list-mode PET was acquired 0 to 45 min after injection. Standard MRI sequences were obtained, including pre- and post-contrast-enhanced (ce) 3-dimensional T1 magnetization prepared for rapid gradient echo imaging (MPRAGE) and 3-dimensional FLAIR. MRSI data parameters included a 2-dimensional PRESS sequence, TR/TE = 1700/30 ms, flip angle of 90^°^, three averages, matrix size of 16 ×16, interpolated to 32×32, vector size of 1024, and a slice thickness of 16 mm, corresponding to a 5.0 × 5.0 × 16 mm^ş^ nominal resolution. The last 15 minutes of the PET/MRI acquisition were used for the reconstruction of static PET images. Image reconstruction was performed with an iterative method (3D OSEM algorithm, 3 iterations, 21 subsets, 344×344 matrix, 4 mm Gaussian filter, voxel size: 2.1× 2.1× 2.0 mm^3^) incorporating point spread function, decay-, scatter-, and attenuation-correction (AC). AC was based on the UTE sequence and the deep learning-based DeepUTE method developed by Ladefoged et al. (48, 49).

### Image and Spectra Analysis

Metabolite maps were generated by analyzing the MR spectrum from each voxel. The MR spectra from brain voxels were processed with phase and frequency corrections before being analyzed using LCModel Version 6.3 (50) over a range of 0.2 to 4.2 ppm, using a basis-set for the Point Resolved Spectroscopy (PRESS) sequence and echo time of 30 ms. This basis set encompassed a wide range of metabolites, including aspartate, Cr, gamma-aminobutyric acid, glutamate, glutamine, glutathione, glycine, glycerophosphocholine (GPC), glycerophosphoethanolamine, Myo-inositol, lactate, NAA, glutamate, phosphocholine (PCh), phosphocreatine (PCr), phosphorylethanolamine, scyllo-inositol, serine, and taurine. For the results, specific metabolite groups were reported as combined measures: tCho as the sum of GPC and PCh, and total creatine (tCr) as the sum of Cr and PCr. To ensure the reliability of the spectral data and metabolite quantification, quality thresholds were applied during LCModel fitting. These criteria included a linewidth of less than 0.15 ppm and a Cramer–Rao lower bound (CRLB) below 25%.

The MRSI-derived metabolite maps were upscaled from 32×32 (in-plane 6.875×6.875 mm^2^) to 128×128 (1.25×1.25 mm^2^), using nearest-neighbor and a trained deep learning model as described previously (51). Custom MATLAB scripts (MathWorks, Natick, MA, USA) were used for the upscaling. The deep learning model is trained to better recover anatomical details during the upscaling compared to the alternative methods, facilitating more accurate localization and quantification of metabolic changes, as described in more detail in the supplementary material. Metabolite ratios, including tCho/NAA and tCho/tCr, were generated and converted into NifTi format. These maps were overlaid on FLAIR and ce-T1 MRI to extract the corresponding voxel values of tCho/NAA and tCho/tCr for each biopsy in FreeView, a viewing platform from FreeSurfer (52). For each biopsy location/voxel, tCho/NAA and tCho/tCr were classified as MRSI positive if the metabolite ratio exceeded the threshold value of 2.0, based on previous reports indicating an abnormally increased level of choline-containing metabolites (15, 53). Based on this threshold criterion, a contour was drawn to include voxels with tCho/NAA and tCho/Cr*≥* 2.0. The MRSI was acquired in a single 2-dimensional slice, covering only parts of the tumor. The number of image-localized biopsies covered by the MRSI slice using the different upscaling methods was therefore evaluated.

Quantitative image analysis in PMOD (version 4.304, PMOD Technologies LLC, Z*ü*rich, Switzerland) involved co-registering ce-T1 MRI and PET to the FLAIR images for each individual. [18F]-FACBC uptake was assessed using a spherical 2 mm radius volume of interest (VOI) at the biopsy sites, to calculate the mean standardized uptake value (SUV) within the VOI. Tumor-to-background ratios (TBRs) were calculated as the mean of the biopsy site (SUV) divided by background uptake defined in the contralateral side of the brain (54). The contra-lateral side of the brain was defined above the ventricles, consisting of six consecutive, crescent-shaped regions of interest, forming a VOI (6). PET volumes were defined using a TBR threshold value of 2.0 (6). PET uptake outside the brain was manually removed. Each biopsy was classified as PET-positive if the biopsy was included in the PET volume (6).

### Statistical Analysis

Statistical analysis was performed in Python using custom scripts. The Fisher exact test was used for categorical variables (e.g., biopsy covered/not covered by MRSI) to evaluate statistical significance. Receiver operating characteristic (ROC) curve analysis assessed the diagnostic accuracy of tCho/NAA, tCho/tCr, TBR, SUV, and combinations of the modalities for classifying glioma vs. non-tumor tissue and IDH1-mutation status. AUC discrimination was classified as follows: <0.5 (none), 0.5–0.7 (poor), 0.7–0.8 (acceptable), 0.8–0.9 (excellent), and ≥0.9 (outstanding) (55). Optimal thresholds were defined as the ROC curve point closest to the top left corner. The DeLong test was used to compare the ROC curves and evaluate statistical differences in their performance. The classification performance of the PET parameters SUV and TBR, and MRSI-derived tCho/NAA and tCho/tCr was investigated as stand-alone parameters, as well as for each combination; e.g., tCho/NAA + SUV, tCho/tCr + TBR. To evaluate the combined classification performance of the combined variables, we fitted a logistic regression model using both variables as predictors. The model estimated predicted probabilities for the outcome, which were then used in an ROC analysis. This approach enabled the assessment of the combined discriminative power of the two variables. We compared the AUC of this combined model to those from the ROC analyses of the individual variables to evaluate improvement in classification performance. When using predicted probabilities from regression in ROC analysis, the thresholds are typically between 0 and 1. These represent decision cutoffs on the probability scale for classifying an observation as positive or negative. We also investigated how accurately these stand-alone and combined parameters classify glioma from non-tumor tissue by calculating their confusion matrices, using the optimal thresholds derived from the ROC analysis.

## Data Availability

Due to ethical and legal restrictions, as well as compliance with the European Union General Data Protection Regulation (GDPR), the dataset acquired and analyzed from St. Olav Hospital and NTNU cannot be made publicly available, as its release would compromise patient privacy. However, investigators may request access to this dataset by contacting Live Eikenes at the Faculty of Medicine and Health Sciences, NTNU, Trondheim. Requests will be subject to a data licensing agreement and institutional approval. Further details on the data transfer agreement process can be found here: (https://i.ntnu.no/wiki/-/wiki/Norsk/Dataoverforingsavtale/. The code for upscaling metabolite maps is available here: https://github.com/MorEsm/SR-MRSI/

## Additional Information and Competing Interests Statement

Authors have no competing interests to declare.

## Acknowledgments

E.B.B. was funded by the Southern Eastern Norway Regional Health Authority (Helse Sør-Øst RHF, HSØ; grant number 2021023).

