## Supplementary Tables 1 & 2 for "Combined High-Resolution MRSI and [18F]-FACBC PET to Improve the Presurgical Diagnostic Accuracy in Gliomas"

**Table S1.** Results from ROC analysis for tCho/NAA, tCho/tCr, SUV, and TBR for classification of glioma versus non-tumor tissue. Sensitivity, specificity, optimal threshold, and AUC with 95% CI are listed. MRSI-derived metabolic ratios were calculated using the deep learning model.

|  | **Deep Learning model** | | | |
| --- | --- | --- | --- | --- |
|  | **Sens.** | **Spes.** | **Threshold** | **AUC**  **(95% CI)** |
| tCho/NAA | 0.79 | 0.88 | 2.37 | 0.87  (0.66-1.00) |
| tCho/tCr | 0.92 | 0.75 | 1.21 | 0.81  (0.54-1.00) |
| SUV | 0.67 | 0.75 | 0.55 | 0.71 (0.49-0.90) |
| TBR | 0.58 | 0.88 | 2.28 | 0.68  (0.48-0.86) |
| tCho/NAA + tCho/tCr | 0.88 | 0.88 | 0.80 | 0.90  (0.69-1.00) |
| tCho/NAA + SUV | 0.83 | 0.88 | 0.76 | 0.89  (0.70-1.00) |
| tCho/NAA + TBR | 0.83 | 0.88 | 0.76 | 0.89  (0.70-1.00) |
| tCho/tCr + SUV | 0.95 | 0.74 | 0.60 | 0.83  (0.59-1.00) |
| tCho/tCr + TBR | 0.96 | 0.75 | 0.61 | 0.83  (0.59-1.00) |
| tCho/NAA + tCho/tCr + SUV | 0.88 | 0.88 | 0.78 | 0.90  (0.69-1.00) |
| tCho/NAA + tCho/tCr + TBR | 0.88 | 0.88 | 0.77 | 0.91  (0.71-1.00) |

**Table S2.** Results from ROC analysis for tCho/NAA, tCho/tCr, SUV, and TBR for classification of *IDH1*-wildtype from *IDH1*-mutated tissue. Sensitivity, specificity, optimal threshold, and AUC with 95% CI are listed.

|  | **Deep Learning model** | | | |
| --- | --- | --- | --- | --- |
|  | **Sens.** | **Spes.** | **Threshold** | **AUC**  **(95% CI)** |
| tCho/NAA | 0.52 | 0.89 | 2.55 | 0.67  (0.44-0.88) |
| tCho/tCr | 0.13 | 1.0 | 2.79 | 0.38  (0.17-0.60) |
| SUV | 0.70 | 1.0 | 0.73 | 0.83  (0.66-0.95) |
| TBR | 0.70 | 0.89 | 2.06 | 0.82  (0.65-0.94) |
| tCho/NAA + tCho/tCr | 0.61 | 0.89 | 0.74 | 0.77  (0.58-0.92) |
| tCho/NAA + SUV | 0.70 | 1.00 | 0.68 | 0.80  (0.62-0.93) |
| tCho/NAA + TBR | 0.70 | 0.89 | 0.69 | 0.83  (0.67-0.96) |
| tCho/tCr + SUV | 1.00 | 0.78 | 0.56 | 0.95  (0.87-1.00) |
| tCho/tCr + TBR | 0.65 | 1.00 | 0.82 | 0.89  (0.76-0.99) |
| tCho/NAA + tCho/tCr + SUV | 1.00 | 0.83 | 0.70 | 0.96  (0.88-1.00) |
| tCho/NAA + tCho/tCr + TBR | 0.70 | 1.00 | 0.77 | 0.90  (0.78-1.00) |
